# Higher Native Peruvian ancestry proportion is associated with tuberculosis progression risk

**DOI:** 10.1101/2020.09.09.20191437

**Authors:** Samira Asgari, Yang Luo, Kamil Slowikowski, Chuan-Chin Huang, Roger Calderon, Judith Jimenez, Rosa Yataco, Carmen Contreras, Jerome T. Galea, Leonid Lecca, David Jones, D. Branch Moody, Megan B. Murray, Soumya Raychaudhuri

## Abstract

The global burden of pulmonary tuberculosis (TB) remains a major public health problem that is particularly severe among and disproportionately affects indigenous populations. We aimed to investigate whether genetic factors related to indigeneity affect TB progression risk in a cohort of admixed Peruvians with active TB and their latently infected household contacts. Our results show that Native Peruvian ancestry is positively associated with TB progression risk: a 10% increase in native ancestry tracks with a 25% increased risk of TB progression. This risk is independent of the potentially confounding socio-demographic and environmental factors that we tested here. Our results demonstrate that the genetic contribution to TB risk varies among populations and brings new insight to the long-standing debate on the role of genetic ancestry in susceptibility to TB. Additionally, our study highlights the value of including diverse populations in genetic studies of infectious diseases and other complex phenotypes, and provides a road map for future similar studies where it is important to account for confounding non-genetic risk factors to identify genetic risk factors.

## Main Text

Tuberculosis (TB), the leading cause of death from an infectious disease worldwide, disproportionately affects indigenous populations^1,2^. Previous^3^ and recent^2,4^ epidemiological studies have firmly established associations between socioeconomic factors such as poverty and TB risk among indigenous populations. The role of genetic factors in the apparently increased susceptibility to TB among indigenous people, on the other hand, has been debated for over 200 years^5,6^. The risk of TB, like that of other infectious diseases, is in part determined by host genetic factors^7–9^. However, there is little concordance between known TB susceptibility loci among different populations^8^, suggesting the possibility that different risk alleles may be driving TB risk in different populations. In line with this observation, epidemiological studies have shown that TB risk differs among populations even after correction for socioeconomic factors^10,11^. It is thus plausible that genetic factors that track with ancestry partially contribute to the high incidence of TB in indigenous populations independently of socioeconomic and environmental factors.

Here we investigate this hypothesis in a prospective cohort of 3,980 Peruvains^9^. The genome of contemporary Peruvians is shaped by extensive admixture between Native Peruvians and the Europeans, Africans, and Asians that arrived in Peru following colonization^12–14^. Our cohort consists of 2,160 HIV-negative patients with microbiologically confirmed pulmonary TB (cases) and 1,820 of their *M. tb* exposed and infected household contacts who had no evidence of pulmonary or extrapulmonary TB (controls, **Figure 1**). Notably, since both cases (TB patients) and controls (latently infected patients) came from the same households, they were exposed to highly similar environmental risk factors. In addition to individuals’ TB status and household information, we have also collected extensive socio-demographic information for each individual(**Table 1**).

**Figure 1:**
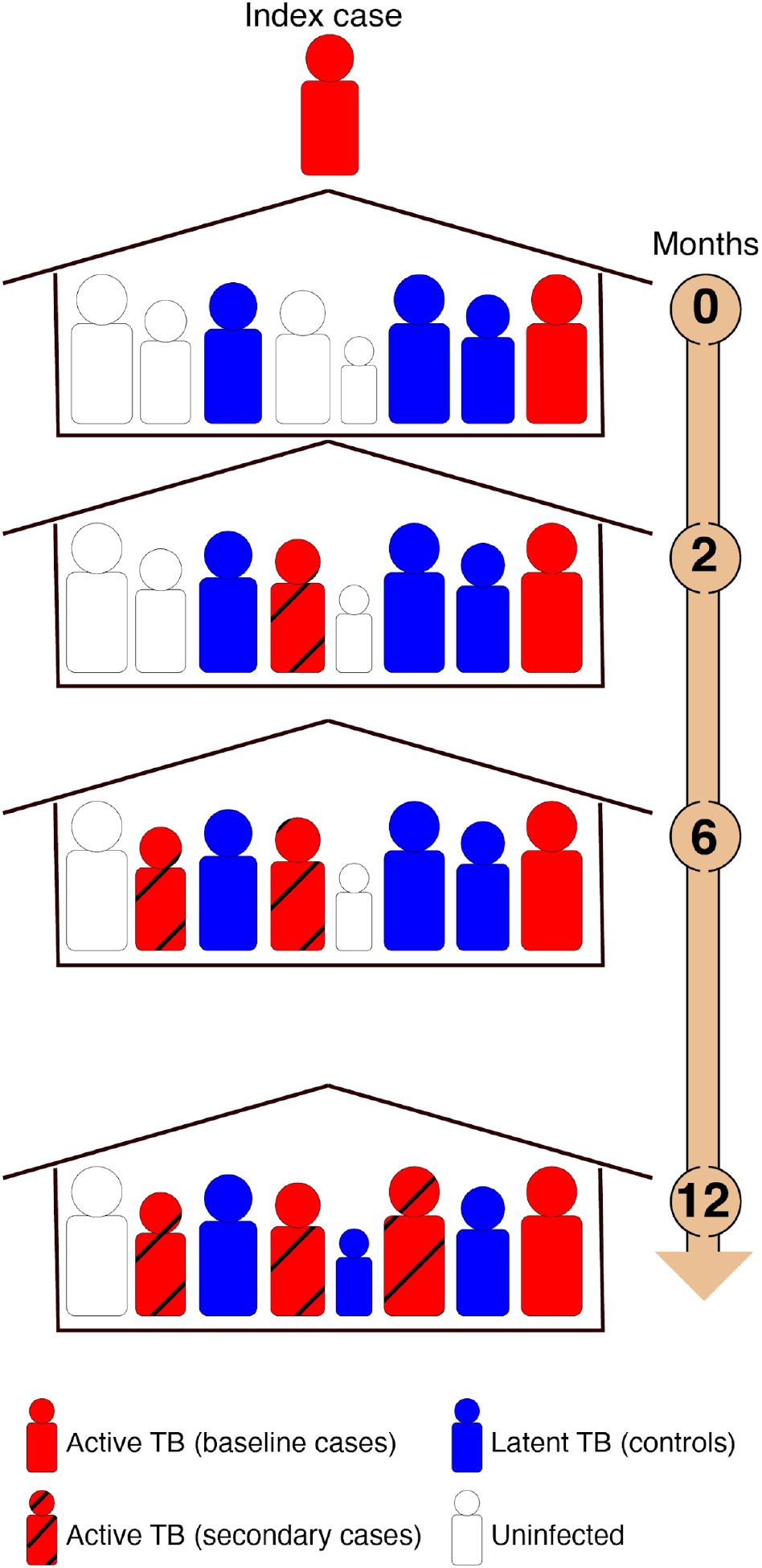
Study design. Patients were recruited in a large catchment area that included 20 urban districts and ∼3.3 million residents. Upon enrollment of index cases (baseline), the household contacts of the index case were contacted within two weeks. Household contacts with pulmonary TB were recruited as cases (baseline cases). Household contacts that were positive for the tuberculin skin test (TST) but did not have active TB were recruited as controls. All individuals were followed up for one year and all household contacts were evaluated for signs and symptoms of pulmonary and extra-pulmonary TB disease at 2, 6, and 12 months after enrollment and were recruited as cases if they developed active TB during follow up (secondary cases). Household contact that remained or became TST positive but did not develop active TB were recruited as controls.

**Table 1:**
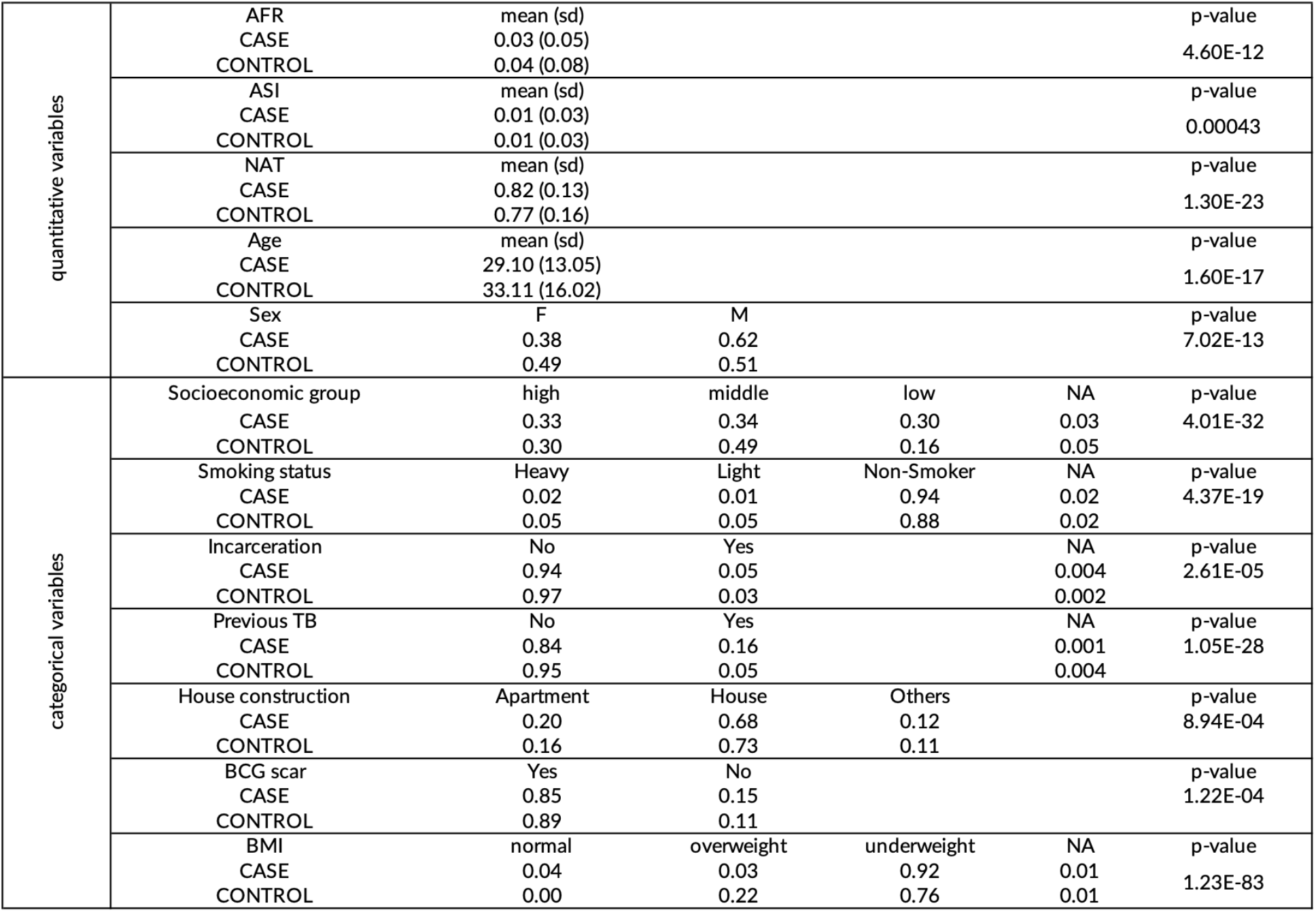
Cohort’s demographic information. The final cohort includes 2,160 individuals with active TB and 1,820 household contacts with latent TB from 2,272 households. Each covariate’s association is tested with TB status individually. P-values are calculated using the t-test for quantitative variables and using the chi-square test for categorical variables. For categorical variables proportions are shown. For quantitative variables, means and standard deviations (sd) are shown. NA: not available.

To investigate the association between Native Peruvian ancestry and TB progression, we first combined our data with genotyping data from the 1000 Genomes Project^15,16^ and from Siberian and Native American populations^17^ (**Methods, Figure S1**). We then inferred global ancestry proportions by performing ADMIXTURE^18^ analysis on the merged dataset (K = 4 clusters, **Methods**). The average proportion of Native Peruvian, European, African, and Asian ancestry was 0.80 (standard deviation (sd) = 0.15), 016 (0.11), 0.03 (0.07), and 0.01 (0.03) respectively (**Figure 2A, Figure S2, Table S1, Table S2**) consistent with previous genetic studies of Peruvians^13,14^. We observed a significantly higher level of Native Peruvian ancestry in cases compared to the household controls (0.82 (0.13) and 0.77 (0.16) respectively, Wilcoxon rank-sum test p-value = 3.36×10^−21^, **Figure 2B**). We performed a series of statistical analyses to determine if this difference was related to population structure, genetic relatedness among the members of the same household, socio-demographic and environmental factors, or factors related to exposure or transmission.

**Figure 2:**
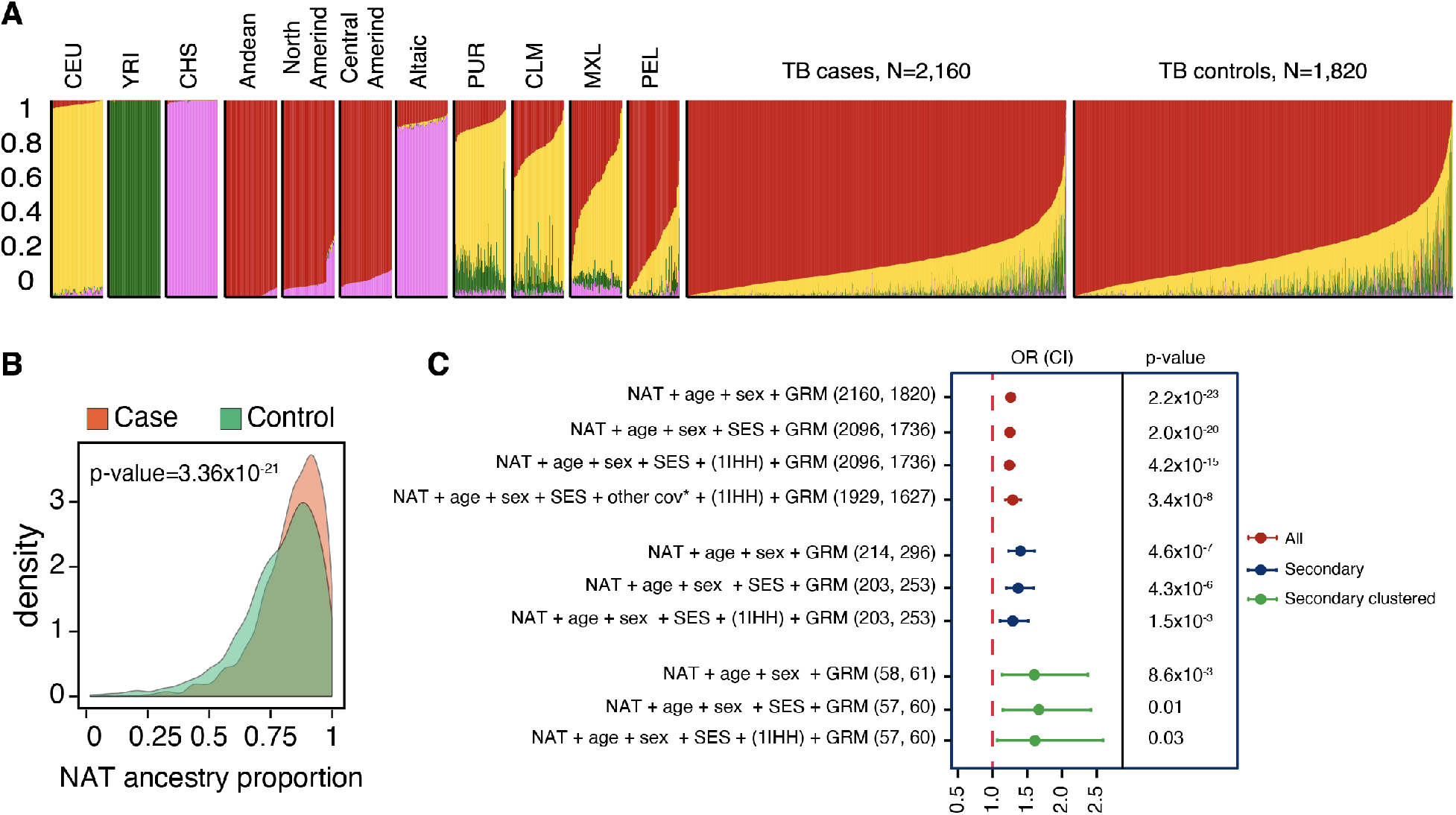
Native Peruvian ancestry is associated with TB progression risk. **A)** Amerind, from Reich et al study), Altaic (Siberians speaking Altaic languages, from Reich et al study), Puerto Ricans from Puerto Rico (PUR from the 1000 Genomes project), Colombians from Medellin, Colombia (CLM from the 1000 Genomes Project), Mexican Ancestry from Los Angeles USA (MXL from the 1000 Genomes Project), Peruvians from Lima, Peru (PEL from the 1000 Genomes Project), TB cases form this study (N = 2160), TB controls form this study (N = 1820). **B)** Probability density distribution of Native Peruvian ancestry proportion in TB cases and controls. TB cases have a higher proportion of Native Peruvian ancestry than household contacts with latent TB (Wilcoxon rank-sum test p-value = 3.36×10^−21^). Y-axis: density. X-axis: Native Peruvian ancestry (NAT) proportion. **C)** Native Peruvian ancestry (NAT) remained significantly associated with TB progression risk after controlling for sex and age as fixed effects and a genetic relatedness matrix (GRM), to account for population structure and genetic relatedness as a random effect, we observed that (p-value = 2.18×10-23, odds ratio (OR) per 0.1 increase in Native Peruvian ancestry proportion (OR_NAT0.1_) = 1.26, 95% confidence interval (CI) = 1.21^−23^). Inclusion of household as a proxy for unmeasured environmental factors as well as additional covariates such as socioeconomic (SES) information and household (HH) did not change this result. Similarly, Native Peruvian ancestry remained significantly associated with TN progression risk when we restricted our cohort to secondary cases (e.g. household contacts who develop TB during the follow up) or secondary clustered cased (e.g secondary cases that their *M. tb* strain shared a molecular fingerprint with the corresponding index case) and their household contacts. circles show odds ratio for 0.1 increase in Native Peruvian ancestry, error bars show 95% CI. *other cov: additional covariates including African and Asian ancestry proportion, smoking, drinking, malnutrition, previous TB history, and incarceration.

Native Peruvian ancestry remained significantly associated with TB progression risk after adjusting for age, sex, and genetic relatedness (base model, chi-square difference test p-value = 2.18×10^−23^, odds ratio per 0.1 increase in Native Peruvian ancestry proportion (OR_NAT0.1_) = 1.26, 95% confidence interval (CI) = 1.21–1.32, **Figure 2C, Table S3**). Increasing the number of ancestral clusters in the ADMIXTURE analysis revealed the finer substructure within each of the four main ancestral populations (Table S2). However, it did not substantively change the association between Native Peruvian ancestry and TB progression risk (**Table S4**).

Next, we adjusted for household-level socioeconomic variables. Data on housing quality, water supply, and sanitation were incorporated into a single continuous variable using principal component analysis^19^ (PCA, **Methods**) which was categorized by tertile to make three socioeconomic status categories. Although socioeconomic status was positively associated with TB progression risk (**Table 1, Table S3**), addition of this variable to our base model had no substantive effect on the strength of the association between Native Peruvian ancestry and TB progression risk (chi-square difference test p-value = 4.14×10^−20^, OR_NAT0.1_ = 1.25 (1.19–1.33), **Figure 2C, Table S3**). We considered the possibility that residual confounders that are not captured by our socioeconomic groups could have biased our association results. To test this, we added a household random effect term to our model positing that individuals from the same household are likely to have similar exposures to exogenous risk factors. Native Peruvian ancestry remained strongly associated with TB progression risk in this model (chi-square difference test p-value = 4.24×10^−15^, OR_NAT0.1_ = 1.24 (1.18–1.31), **Figure 2C, Table S3**).

To address the possibility that Native Peruvian ancestry acts as a proxy for other socio-demographic factors collected in our cohort that are also associated with TB risk (**Table 1**), in addition to covariates described above, we included the following covariates in our model: African and Asian ancestry proportion, smoking, alcohol use, BMI, previous TB history, and incarceration in our model (**Table 1**). Similar to household and socioeconomic status, the inclusion of these covariates did not obviate the significance or reduce the effect size (chi-square difference test p-value = 1.09×10^−6^, OR_NAT0.1_ = 1.26 (1.15–1.38), **Figure 2C, Table S3**). Collectively these results suggest that relative to European ancestry, Native Peruvian ancestry is associated with increased TB progression risk independent of socio-demographic factors and population structure that were tested here.

In our cohort, index cases are more likely to have acquired TB in the community, while household contacts who developed TB after the diagnosis of index cases (secondary cases) are more likely to result from within household transmission. We considered whether the apparent ancestry differences could be explained by potential differences in exposure and transmission between the index case and household contacts. To address this, we performed an analysis which included only secondary cases (N = 214) and their household contacts who did not develop TB during 1 year of follow up (N = 296, **Methods, Figure 1**). In this analysis we observed an odds ratio similar to the one observed for the whole cohort (using the base model OR_NAT0.1_ = 1.40 (1.23–1.61), chi-square difference test p-value = 4.5×10^−5^, **Table S3, Figure 2C**). We further restricted the cohort to secondary TB cases who shared an *M. tb* molecular fingerprint with the corresponding index case to ensure that their TB disease was the result of within household transmission rather than infection from other community sources or reactivation of a previous infection. While this analysis was much smaller, we observed that it did not substantively change the association results (N = 56 TB cases and 61 household contacts, using the base model OR_NAT0.1_ = 1.58 (1.12–2.23), chi-square difference test p-value = 8.6×10^−3^, **Table S3, Figure 2C
**).

Collectively, these analyses bring additional support for our primary finding and alleviate the key concern that the association between Native Peruvian ancestry and TB progression risk is due to apparent biases in exposure risk between cases and controls. In line with this conclusion, we did not find significant association between Native Peruvian ancestry and TB progression risk when we compared baseline TB cases with secondary cases from the same households (N = 138 and 214 respectively, using the base model OR_NAT0.1_ = 1.05 (0.89–1.24), chi-square difference test pvalue = 0.54, **Table S3**) showing that differences in exposure do not explain differences in Native Peruvian ancestry proportion.

To identify specific genomic regions that might explain the association between Native Peruvian ancestry and TB progression risk, we performed local ancestry inference followed by admixture mapping in our cohort using PCAdmix^20^ and reference populations from the 1000 Genomes Project^21^ and the Simons Genome Diversity Project^22^ (**Methods**). We then used the local ancestry inference results to search for genomic regions with significantly higher Native Peruvian ancestry in TB cases compared to household contacts after correction for age, sex, global ancestry proportions, and genetic relatedness (**Methods**). We used permutation to determine a genome-wide significance threshold of 1.09×10^−5^ for admixture mapping in Peruvians which is similar to the previously suggested p-value threshold 5.7×10^−5^ for admixture mapping in Hispanic/Latino populations^23^. No individual locus passed this stringent significance threshold. We observed nominal evidence of association at 3q23 (Wald test Wald test p-value = 2.8×10^−5^), 2p24.3 (Wald test p-value = 5.9×10^−5^), and 5p23.2 (Wald test p-value = 7.2×10^−5^, **Figure S3**). In a previous genome-wide association study (GWAS) our group showed that 3q23 locus is significantly associated with TB progression risk in Peruvians (p-value < 5×10^−8^)^9^. In our GWAS, the top risk variant (rs73226617, OR = 1.18; p-value = 3.9×10^−8^) at 3q23 has a higher frequency in Europeans compared to Peruvians (MAF = 0.05 vs. 0.03) and is unlikely to explain the admixture mapping signal we observe at this locus. Nonetheless, the overlap between GWAS and admixture mapping signal suggests the potential role of genes and variants overlapping 3q23 in TB progression risk in Peruvians.

TB progression is a complex trait with an estimated SNP heritability (h^2^
_g_) of 21.2% (standard error = 0.08)^9^ in Peruvians. However, previous GWAS of TB susceptibility and TB progression identified only a handful of variants with relatively small effect sizes which explain only a small proportion of TB progression heritability suggesting that more risk variants are yet to be identified. Similarly, the absence of any single locus achieving statistical significance in our admixture mapping suggests that TB progression is driven by a polygenic architecture where many variants with modest effect sizes are driving the effect of Native Peruvian ancestry on TB progression risk. Conclusively identifying such loci requires larger studies with greater statistical power.

There is little concordance between TB risk loci identified through GWAS in different populations^9^, suggesting different alleles may contribute to TB risk in different populations. This hypothesis is supported by our results which show that TB progression risk differs among populations with different genetic ancestries. In line with our findings, a previous study of TB risk in the admixed South African Coloured (SAC) population identified an association between San ancestry and susceptibility to TB^10^. Similarly, associations between genetic ancestry and infectious disease outcomes have been reported for other infections such as cholera^24^. This is not surprising, because pathogens are among the strongest selective forces in human evolution^25,26^. In the case of TB specifically, there is growing evidence that *M. tb* and the human genome have co-evolved^27^. While *M. tb* has been present in Europe for several thousands of years^28^, the overall consensus is that it arrived in Peru much more recently and following colonization^29^. Although archeological evidence suggests that mycobacterial diseases existed in Peru before colonization^30,31^, whole-genome sequencing of samples from pre-Columbian mummies in Peru suggested *Mycobacterium pinipeneddi* or other mycobacteria as the causal pathogen for these diseases^31^. Moreover, currently circulating *M. tb* strains in Peru as well as the rest of the Americas are largely dominated by the Euro-American lineage 4 genotypes^32,33^ supporting the hypothesis that modern strains of *M. tb* were brought to Peru through the European colonization. It is thus possible that the long, shared history of Europeans and TB has led to selective pressures that have mitigated TB risk, and that such pressures have not been present in the Americas. Increased resistance to a specific pathogen due to co-evolution has been reported before and in other populations. For example, analysis of positively selected genomic loci in a population from the Ganges River Delta, the historic geographic epicenter of cholera, showed selection for genes involved in immunity to *Vibrio cholerae* in this population^24^.

Besides exposure to *M. tb*, it is plausible that differences in exposure to other pathogens account for the population differences in the risk of TB progression observed in our study. The idea that differential exposure to pathogens over historical time could have led to differential susceptibility to different pathogens among populations is an old one^34,35^. Modern evidence for this idea comes from studies in mouse models that have shown that microbial exposure can increase resistance to bacterial and parasitic pathogens by affecting the basal immune system and the composition of the innate and adaptive immune cells^36,37^ suggesting that differences in exposure history can lead to differences in the immune system. It is possible that exposure to pathogens that were highly endemic in the old world populations and were absent in the pre-colonial Peru^38,39^ has led to increased resistance to *M. tb* and possibly other pathogens in Europeans. However, such arguments must be made with great care as discussed in detail by previous researchers^40^, and as we have worked to do here.

One caveat of our study is that we have not tested for all possible non-genetic TB risk factors. For example, while we tried to account for factors related to exposure by performing analysis on secondary cases we do not have information on community and workplace exposures, nor do we have access to childhood exposure history to other pathogens that might have mitigated TB risk. In our study we have controlled for factors related to pathogen exposure and transmission by restricting our analyses to secondary cases who shared an *M. tb* molecular fingerprint with the corresponding index case. However, we have not checked for bacterial strain heterogeneity, and cannot comment on the possible differences in pathogenicity of specific *M. tb* strains among ancestries. It is important to note that our results cannot be generalized to all indigenous populations as different populations have different population histories and the socio-demographic factors, which are the primary determinants of TB risk, vary widely across different indigenous populations^2,4^.

In addition to its relevance for better understanding the genetic architecture of TB progression, our study highlights the challenges associated with disentangling the role of heritable ancestry-specific genetic factors from the non-heritable factors that correlate with ancestry^41,42^ and provides a framework for future similar studies where it is important to account for environmental/socioeconomic factors to identify genetic factors that affect disease outcome. Specifically, such studies need to be large enough to be statistically well-powered; they need to control for potential socioeconomic and environmental confounders^2^, including those that cannot environmental/socioeconomic factors to identify genetic factors that affect disease outcome be easily measured such as exposure or transmission; ancestry needs to be genetically defined because self-reported ancestry is often inaccurate^43,44^; and the disease phenotypes need to be precisely defined as different genetic factors might underlie under different stages of the disease (e.g infection versus progression upon infection)^9^.

Finally, our results highlight that differences in infectious disease burden among different populations cannot be solely attributed to variations in sociodemographic factors and underline the importance of conducting large scale host genetic studies in diverse populations in order to reduce health disparities and to get a comprehensive picture of the genotype-phenotype relationship in TB and other infectious diseases.

## Material and Methods

### Study design & participants

As described in detail elsewhere^45^, the study was conducted in 106 district health centers in Lima, Peru in a catchment area including 12 of the 43 districts of metropolitan Lima and 3.3 million inhabitants. Samples were obtained following institutional IRB guidelines and with informed consent from participants. Pulmonary TB patients were diagnosed by the presence of acid-fast bacilli in sputum smear or a positive *M. tb* culture at any time from enrollment to the end of treatment as described^45^. Household contacts were enrolled within two weeks of enrolling an index case at which time their infection status was determined using the Tuberculin Skin Test (TST). Household contacts were evaluated for signs and symptoms of pulmonary and extrapulmonary TB disease at two, six, and 12 months after enrollment. Those that remained TST positive and did not develop active TB during the follow-up period were included as controls in the study. Household contacts that developed active TB 14 days or more after index case enrollment were considered secondary cases. For those with positive TB cultures, isolates underwent drug-susceptibility testing (DST) and MIRU-based genotyping.

Besides individuals’ TB status, we also collected extensive information for each individual^45^ including age, gender, socio-demographic factors, BCG vaccination, alcohol and tobacco use, comorbid disease, and previous TB disease. We categorized participants according to their alcohol intake as follows: nondrinkers if they reported having consumed no alcoholic drinks per day, light drinkers if they reported drinking < 40 grams or < 3 alcoholic drinks per day and heavy drinkers if they reported drinking 40 grams of alcohol or more or 3 or more drinks per day^45^. For smoking, we classified people as nonsmokers if they reported no cigarette smoking, as light smokers if they reported smoking one cigarette per day and as heavy smokers if they reported smoking more than one cigarette per day^45^. We categorized people with BMI *z-*scores of less than 2 as underweight and those greater than 2 as overweight. We defined a nutritional status for children based on the World Health Organization body mass index (BMI) z-score tables^46^.

In addition to individual-level data, we collected household-level socioeconomic factors on housing quality, water supply, and sanitation^19^. We calculated household-level composite socioeconomic scores (SES scores) by summarizing these factors using principal component analysis (PCA)^19^. The continuous SES scores were categorized into tertiles corresponding to low, middle, and upper socioeconomic groups.

### Genotyping and quality control

As we have described in detail elsewhere^9^, genomic DNA was extracted from participants’ whole blood. All samples were genotyped using a 720K customized Affymetrix Axiom array. Quality control on the genotypes, phasing, and imputation was performed as described previously^9^. In brief, we excluded duplicated individuals, individuals with genotype missing rate > 5%, and individuals with an excess of heterozygous genotypes (3.5 standard deviations from the mean). We then excluded duplicated variants, variants with missingness rate > 5%, variants with a batch effect (p-value < 10^−5^), variants with Hardy-Weinberg equilibrium (HWE) p-value < 10^−5^ in controls, and a missing rate per SNP difference in cases and controls > 10^−5^. The final cohort included 677,232 variants and 3,980 individuals including 2,160 HIV-negative, culture-positive, drug-sensitive TB cases, and 1820 household contacts.

### Principal Component Analysis (PCA) and global ancestry inference

The genome of admixed individuals is a mosaic of chromosomal blocks from different ancestral populations. The average proportion of these blocks from each ancestral population over the whole genome is a measure per individual admixture levels and is referred to as “global ancestry proportion”^47^. We merged genotyping data from our cohort, with previously published data from the 1000 Genomes Project phase 3 (2,054 individuals from 26 populations)^15,16^ and Siberian and Native American populations from Reich et *al*.^17^(493 individuals from 57 Native American populations and 245 individuals from 17 Siberian populations)^17^, by matching on the chromosome, position, reference, and alternate alleles using PLINK (version 1.90b3w, https://www.coggenomics.org/plink/1.9/)^48^. After merging the datasets, variants with an overall MAF < 1% were excluded. The final merged dataset included 34,936 variants. We used the Genome-wide Complex Trait Analysis tool (GCTA^49^, version 1.26.0) to perform PCA on this dataset. To estimate the global ancestry proportions, we pruned the data for linkage disequilibrium by removing the markers with r^2^ > 0.1 with any other marker within a sliding window of 50 markers per window and an offset of 10 using PLINK (version 1.90b3w). We then performed unsupervised clustering using ADMIXTURE^18^ (version 1.3) with K = 3–7 clusters on the merged and pruned dataset (N = 22,266 variants) to infer the global ancestry proportions. The identity of resulting ancestry proportions in admixed Peruvians was defined based on comparison with ancestry proportions of reference populations.

### Kinship estimation

Many kinship estimation methods perform under the assumption of sampling from a single population with no underlying ancestral diversity. Kinship estimates are inflated when this assumption is violated^50^. In the presence of population structure and admixture, methods that replace population allele frequencies with ancestry-specific allele frequencies are preferred^50^. We used GENESIS R package (version 2.6.1) to estimate the kinship coefficients between individuals. The package is based on the PC-Relate^51^ method, and estimates the individual-specific allele frequencies using linear predictors of top principal components. We considered individuals unrelated if their estimated kinship coefficients were ≤ 0.0625, corresponding to second-degree genetic relatedness or more distant relatives. 468 individuals had kinship coefficients > 0.0625 (e.g were related). Genomic regions with long-range linkage disequilibrium (LD)^52^ including the major histocompatibility complex were excluded for kinship estimation.

### Genetic relatedness matrix (GRM)

To avoid spurious association results, we accounted for both recent genetic relatedness, such as family structure, and more distant genetic relatedness, such as population structure. To generate a genetic relatedness matrix (GRM), we removed rare variants (MAF ≤ 1%), regions with known long-range linkage disequilibrium (LD)^52^, and variants in high LD (r^2^ > 0.2 in a window of 50kb and an offset of 5) using Plink (version 1.90b3w). We then used the Genome-wide Efficient Mixed Model Association (GEMMA^53^, version 0.96) software, with default options, to calculate the GRM. Genomic regions with long-range LD^52^ including the major histocompatibility complex were excluded for GRM calculation.

### Correlation between global ancestry proportions and TB progression risk

We used the R package lme4qtl^54^, a linear mixed model framework, to measure the association between Native Peruvian ancestry proportion and TB progression risk. We included the following covariates in the *base model*: age, sex, as well as a GRM generated using GEMAA^53^ to account for population structure and genetic relatedness. We repeated this analysis after 1) adding African and Asian ancestry proportions; 2) adding household-level socioeconomic groups; 2) adding of a random effect to account for individual’s household; and 3) adding smoking status, drinking status, education, isoniazid preventive therapy, BCG vaccination status, malnutrition, previous TB, and incarceration as covariates.

To further control for factors related to *M. tb* transmission we performed two sets of additional analyses. First, we restricted the analysis to 214 secondary cases and their household contacts(N = 296) and measured the association between Native Peruvian ancestry proportion and TB progression risk using the base model as well as model 1 and 2. Then, we further restricted the cases to secondary cases whose *M. tb* strains shared an identical or similar MIRU pattern (i.e. shared molecular fingerprint) with an index patient (N = 58) and their household contacts (N = 61) and measured the association between Native Peruvian ancestry proportion and TB progression risk using the base model as well as model 1 and 2. For these two sets of analyses, we did not use model 3 as a large number of covariates combined with the small number of samples prevented the models from converging.

For all above analyses, we used the chi-square difference test to compare nested models with or without a given covariate.

### Local ancestry inference

“Local ancestry” is defined as the genetic ancestry of an individual at a particular locus, where an individual can have 0, 1, or 2 copies of an allele derived from each ancestral population^55^. We performed local ancestry inference using PCAdmix and a new reference panel for Native Ancestry, in this analysis we used imputed data to increase the number of shared markers between our data and reference data. Following imputation as described previously^9^, we excluded SNPs with imputation quality score *r*^2^ < 0.4, HWE p-value < 10^−5^ in controls, or a missing rate per SNP greater than 5% which left 7,756,401 markers. We used the Native American individuals from the Simons Genome diversity project^22^ (N = 25 individuals) as well as 5 individuals form the 1000 Genomes Project^21^ PEL that had inferred Native ancestry > 0.95 based on ADMIXTURE analysis at K = 4 clusters as a proxy for Native Peruvian ancestry (N = 30 individuals in total). We used 30 randomly selected individuals from CEU, 30 randomly selected individuals from YRI, 30 randomly selected individuals from CHS populations in the 1000 Genomes study as proxies for European, African, and Asian ancestries respectively. We then merged our data with the reference panel data and restricted the merged dataset to variants with MAF > = 5% in each of the reference populations. We phased the merged data using SHAPEIT as described above. After phasing and following PCAdmix developer’s recommendation, we used PLINK (version 1.90b3w) to remove the markers with r^2^ > 0.8 with any other marker within a sliding window of 20 markers per window and an offset of 10 using. We then used SHAPEIT2^56^ (version v2.r837) to generate VCF files followed by Beagle (version 4.1) to generate input files for PCAdmix. All files were generated per chromosome. Finally, we performed local ancestry inference on each chromosome using PCAdmix (version 3) with the following option –bed and –ld 0 and recombination maps from the 1000 Genomes Project^21^. Local ancestry inference was done in windows of 20 SNPs and in total local ancestry was inferred for 44,470 intervals. Genomic regions with long-range LD^52^ including the major histocompatibility complex were excluded for admixture mapping.

### Admixture mapping

We used admixture mapping, a method to associate the ancestry of a locus with a trait in an admixed population^57^, to search for genomic loci that can explain some of the observed association between Native Peruvian ancestry and height. We used the Generalized Mixed Model Association Test (GMMAT^58^, version 1.0.3), a generalized linear mixed model framework to check the association between the inferred local Native Peruvian ancestry {coded as a number between 0 and 2 for local Native ancestry posterior probability, 0 means no NAT allele and 2 means 2 NAT alleles} and TB progression risk {case, control}. Standardized age {0 ≤ age ≤ 1}, sex {male, not-male}, global African (AFR) and European (EUR) ancestry proportions {0 ≤ AFR/EUR ≤ 1}, and genetic relatedness (GR) {0 ≤ GR ≤ 0.5} calculated using PC-Relate^51^ were included as covariates in the model. To define the significance threshold for admixture mapping we permuted the case-control status and repeated the association analysis 1000 times. We then used the lowest p-value from each permutation to generate an empirical null distribution. The fifth percentile of this distribution was used as the cutoff for genome-wide significance. The significance threshold we found using this approach was 1.09×10^−5^ which is similar to the previously suggested *p*-value threshold 5.68×10^−5^ for admixture mapping in Hispanic/Latino populations^23^. Total number of loci tested was 44,470.

## Data Availability

Individual-level phenotypes and genotyping data are available through dbGAP, under accession number phs002025.v1.p1.

https://www.ncbi.nlm.nih.gov/projects/gap/cgi-bin/study.cgi?study_id=phs002025.v1.p1

## Acknowledgement

The study was supported by the National Institutes of Health (NIH) TB Research Unit Network, grants U19-AI111224–01 and U01-HG009088 and NIH grants U01-HG009379, and 1R01AR063759. The content is solely the responsibility of the authors and does not necessarily represent the official views of the NIH. S.A. was supported by the Swiss National Science Foundation (SNSF) postdoctoral mobility fellowships P2ELP3_172101 and P400PB_183823.

## Code availability

No custom code was used to draw the central conclusions of this work. All the software and packages used in this work are included and referenced in the manuscript.

## Notes

### Competing Interest Statement

The authors have declared no competing interest.

### Author Declarations

We obtained written informed consent from all the participants. The study protocol was approved by the Institutional Review Board of Harvard School of Public Health and by the Research Ethics Committee of the National Institute of Health of Peru.

## References

1. Tollefson, D. et al. Burden of tuberculosis in indigenous peoples globally: a systematic review. Int. J. Tuberc. Lung Dis. 17, 1139–1150 (2013).

2. Cormier, M. et al. Proximate determinants of tuberculosis in Indigenous peoples worldwide: a systematic review. Lancet Glob Health 7, e68–e80 (2019).

3. National Tuberculosis Association. Committee on tuberculosis among the North American Indians. Tuberculosis Among the North American Indians: Report of a Committee of the National Tuberculosis Association Appointed on October 28, 1921, on Tuberculosi. Among the North American Indians. (U.S. Government Printing Office, 1923).

4. Basta, P. C. & de Sousa Viana, P. V. Determinants of tuberculosis in Indigenous people worldwide. The Lancet. Global health vol. 7 e6–e7 (2019).

5. McMillen, C. W. ‘The Red Man and the White Plague’: Rethinking Race Tuberculosis, and American Indians ca. 1890–1950. Bull. Hist. Med. 82, 608–645 (2008).

6. Jones, D. S. Virgin Soils Revisited. William Mary Q. 60, 703–742 (2003).

7. Naranbhai, V. The Role of Host Genetics (and Genomics) in Tuberculosis. Microbiol Spectr 4, (2016).

8. Abel, L. et al. Genetics of human susceptibility to active and latent tuberculosis: present knowledge and future perspectives. Lancet Infect. Dis. 18, e64–e75 (2018).

9. Luo, Y. et al. Early progression to active tuberculosis is a highly heritable trait driven by 3q23 in Peruvians. Nat. Commun. 10, 3765 (2019).

10. Chimusa, E. R. et al. Genome-wide association study of ancestry-specific TB risk in the South African Coloured population. Hum. Mol. Genet. 23, 796–809 (2014).

11. Daya, M. et al. Using multi-way admixture mapping to elucidate TB susceptibility in the South African Coloured population. BMC Genomics 15, 1021 (2014).

12. Harris, D. N. et al. Evolutionary genomic dynamics of Peruvians before, during, and after the Inca Empire. Proceedings of the National Academy of Sciences 115, E6526–E6535 (2018).

13. Homburger, J. R. et al. Genomic Insights into the Ancestry and Demographic History of South America. PLoS Genet. 11, e1005602 (2015).

14. Ruiz-Linares, A. et al. Admixture in Latin America: geographic structure, phenotypic diversity and self-perception of ancestry based on 7,342 individuals. PLoS Genet. 10, e1004572 (2014).

15. Consortium, 1000 et al. A global reference for human genetic variation. Nature 526, 68–74 (2015).

16. Sudmant, P. H. et al. An integrated map of structural variation in 2,504 human genomes. Nature 526, 75–81 (2015).

17. Reich, D. et al. Reconstructing Native American population history. Nature 488, 370–374 (2012).

18. Alexander, D., Novembre, J. & Lange, K. Fast model-based estimation of ancestry in unrelated individuals. Biotechfor 19, 1655–1664 (2009).

19. Odone, A. et al. Acquired and Transmitted Multidrug Resistant Tuberculosis: The Role of Social Determinants. PLoS One 11, e0146642 (2016).

20. Brisbin, A. et al. PCAdmix: principal components-based assignment of ancestry along each chromosome in individuals with admixed ancestry from two or more populations. Hum. Biol. 84, 343–364 (2012).

21. Abecasis, G. R. et al. An integrated map of genetic variation from 1,092 human genomes. Nature 491, 56–65 (2012).

22. Mallick, S. et al. The Simons Genome Diversity Project: 300 genomes from 142 diverse populations Nature 538, 201–206 (2016).

23. Brown, L. A. et al. Admixture Mapping Identifies an Amerindian Ancestry Locus Associated with Albuminuria in Hispanics in the United States. J. Am. Soc. Nephrol. 28, 2211–2220 (2017).

24. Ryan, E. T., Qadri, F. & Sabeti, P. C. Natural selection in a bangladeshi population from the cholera-endemic ganges river delta. Sci. Transl. Med. (2013).

25. Karlsson, E. K., Kwiatkowski, D. P. & Sabeti, P. C. Natural selection and infectious disease in human populations. Nat. Rev. Genet. 15, 379–393 (2014).

26. Barreiro, L. B. & Quintana-Murci, L. From evolutionary genetics to human immunology: how selection shapes host defence genes. Nat. Rev. Genet. 11, 17–30 (2010).

27. Brites, D. & Gagneux, S. Co-evolution of M ycobacterium tuberculosis and H omo sapiens. Immunol. Rev. 264, 6–24 (2015).

28. Comas, I. et al. Out-of-Africa migration and Neolithic coexpansion of Mycobacterium tuberculosis with modern humans. Nat. Genet. 45, 1176–1182 (2013).

29. Woodman, M., Haeusler, I. L. & Grandjean, L. Tuberculosis Genetic Epidemiology: A Latin American Perspective. Genes 10, (2019).

30. Mackowiak, P. A., Blos, V. T., Aguilar, M. & Buikstra, J. E. On the origin of American tuberculosis. Clin. Infect. Dis. 41, 515–518 (2005).

31. Bos, K. I. et al. Pre-Columbian mycobacterial genomes reveal seals as a source of New World human tuberculosis. Nature 514, 494–497 (2014).

32. Grandjean, L. et al. The Association between Mycobacterium Tuberculosis Genotype and Drug Resistance in Peru. PLoS One 10, e0126271 (2015).

33. Wiens, K. E. et al. Global variation in bacterial strains that cause tuberculosis disease: a systematic review and meta-analysis. BMC Med. 16, 196 (2018).

34. William, M. & Others. Plagues and peoples. (1976).

35. Crosby, A. W. Jr. The Columbian Exchange: Biological and Cultural Consequences of 1492, 30th Anniversary Edition. (ABC-CLIO, 2003).

36. Beura, L. K. et al. Normalizing the environment recapitulates adult human immune traits in laboratory mice. Nature 532, 512–516 (2016).

37. Tamburini, S., Shen, N., Wu, H. C. & Clemente, J. C. The microbiome in early life: implications for health outcomes. Nat. Med. 22, 713–722 (2016).

38. Sessa, R., Palagiano, C., Scifoni, M. G., di Pietro, M. & Del Pian, M., The major epidemic infections: a gift from the Old World to the New? Panminerva Med. 41, 78–84 (1999).

39. Diamond, J. M. Guns, Germs and Steel: A Short History of Everybody for the Last 13,000 Years. (Vintage, 1998).

40. Cameron, C. M., Kelton, P. & Swedlund, A. C. Beyond Germs: Native Depopulation in North America. (University of Arizona Press, 2015).

41. Williams, D. R., Priest, N. & Anderson, N. B. Understanding associations among race, socioeconomic status, and health: Patterns and prospects. Health Psychol. 35, 407–411 (2016).

42. Williams, D. R., Mohammed, S. A., Leavell, J. & Collins, C. Race, socioeconomic status, and health: complexities, ongoing challenges, and research opportunities. Ann. N. Y. Acad. Sci. 1186, 69–101 (2010).

43. Klimentidis, Y. C., Miller, G. F. & Shriver, M. D. admixture, self-reported ethnicity, self-estimated admixture, and skin pigmentation among Hispanics and Native Americans. American Journal of Physical Anthropology: The Official Publication of the American Association of Physical Anthropologists 138, 375–383 (2009).

44. Kumar, R. et al. Genetic ancestry in lung-function predictions. N. Engl. J. Med. 363, 321–330 (2010).

45. Becerra, M. C. et al. Resistance at No Cost: The Transmissibility and Potential for Disease Progression of Drug-Resistant M. Tuberculosis. bioRxiv 475764 (2018) doi:10.1101/475764.

46. de Onis, M. & BlöBlossner, M. The World Health Organization Global Database on Child Growth and Malnutrition: methodology and applications. International Journal of Epidemiology vol. 32 518–526 (2003).

47. Tang, H., Peng, J., Wang, P. & Risch, N. J. Estimation of individual admixture: analytical and study design considerations. Genet. Epidemiol. 28, 289–301 (2005).

48. Chang, C. C. et al. Second-generation PLINK: rising to the challenge of larger and richer datasets. Gigascience 4, 7 (2015).

49. Yang, J., Lee, S. H., Goddard, M. E. & Visscher, P. M. GCTA: a tool for genome-wide complex trait analysis. Am. J. Hum. Genet. 88, 76–82 (2011).

50. Manichaikul, A. et al. Robust relationship inference in genome-wide association studies. Bioinformatics 26, 2867–2873 (2010).

51. Conomos, M. P., Reiner, A. P., Weir, B. S. & Thornton, T. A. Model-free Estimation of Recent Genetic Relatedness. Am. J. Hum. Genet. 98, 127–148 (2016).

52. Price, A. L. et al. Long-range LD can confound genome scans in admixed populations. American journal of human genetics vol. 83 132–5; author reply 135–9 (2008).

53. Zhou, X. & Stephens, M.Efficient multivariate linear mixed model algorithms for genome-wide association studies. Nat. Methods 11, 407–409 (2014).

54. Ziyatdinov, A. et al. lme4qtl: linear mixed models with flexible covariance structure for genetic studies of related individuals. BMC Bioinformatics 19, 68 (2018).

55. Thornton, T. A. & Bermejo, J. L. Local and global ancestry inference and applications to genetic association analysis for admixed populations. Genet. Epidemiol. **38 Suppl 1**, S5–S12 (2014).

56. O’Connell, J. et al. A general approach for haplotype phasing across the full spectrum of relatedness. PLoS Genet. 10, e1004234 (2014).

57. Patterson, N. et al. Methods for high-density admixture mapping of disease genes. Am. J. Hum. Genet. 74, 979–1000 (2004).

58. Chen, H. et al. Control for Population Structure and Relatedness for Binary Traits in Genetic Association Studies via Logistic Mixed Models. Am. J. Hum. Genet. 98, 653–666 (2016).

